# Ethnic differences in the indirect impacts of the COVID-19 pandemic on clinical monitoring and hospitalisations for non-COVID conditions in England: An observational cohort study using OpenSAFELY

**DOI:** 10.1101/2023.01.04.23284174

**Authors:** Ruth E Costello, John Tazare, Dominik Piehlmaier, Emily Herrett, Edward PK Parker, Bang Zheng, Kathryn E Mansfield, Alasdair D Henderson, Helena Carreira, Patrick Bidulka, Angel YS Wong, Charlotte Warren-Gash, Joseph F Hayes, Jennifer K Quint, Brian MacKenna, Rosalind M Eggo, Srinivasa Vittal Katikireddi, Laurie Tomlinson, Sinéad M Langan, Rohini Mathur, the longitudinal health and wellbeing collaborative and the OpenSAFELYcollaborative

## Abstract

**Background:** The COVID-19 pandemic disrupted healthcare and may have impacted ethnic inequalities in healthcare. We aimed to describe the impact of pandemic-related disruption on ethnic differences in clinical monitoring and hospital admissions for non-COVID conditions in England.

**Methods:** We conducted a cohort study using OpenSAFELY (2018-2022). We grouped ethnicity (exposure), into five categories: White, South Asian, Black, Other, Mixed. We used interrupted time-series regression to estimate ethnic differences in clinical monitoring frequency (e.g., blood pressure measurements) before and after 23rd March 2020. We used multivariable Cox regression to quantify ethnic differences in hospitalisations related to: diabetes, cardiovascular disease, respiratory disease, and mental health before and after 23rd March 2020.

**Findings:** Of 14,930,356 adults in 2020 with known ethnicity (92% of sample): 86.6% were White, 7.3% Asian, 2.6% Black, 1.4% Mixed ethnicity, and 2.2% Other ethnicities. Clinical monitoring did not return to pre-pandemic levels for any ethnic group. Ethnic differences were apparent pre-pandemic, except for diabetes monitoring, and remained unchanged, except for blood pressure monitoring in those with mental health conditions where differences narrowed during the pandemic. For those of Black ethnicity, there were seven additional admissions for diabetic ketoacidosis per month during the pandemic, and relative ethnic differences narrowed during the pandemic compared to White. There was increased admissions for heart failure during the pandemic for all ethnic groups, though highest in White ethnicity. Relatively, ethnic differences narrowed for heart failure admission in those of Asian and Black ethnicity compared to White. For other outcomes the pandemic had minimal impact on ethnic differences.

**Interpretation:** Our study suggests ethnic differences in clinical monitoring and hospitalisations remained largely unchanged during the pandemic for most conditions. Key exceptions were hospitalisations for diabetic ketoacidosis and heart failure, which warrant further investigation to understand the causes.

**Funding:** LSHTM COVID-19 Response Grant (DONAT15912).

**Research in context:** *Evidence before this study:* We searched MEDLINE from inception to 7th September 2022, for articles published in English, including the title/abstract search terms (healthcare disruption OR indirect impact OR miss* diagnos* OR delayed diagnos* OR service disruption) AND (sars-cov-2 OR covid-19 OR pandemic OR lockdown) AND (ethnic*). Of the seven studies identified, two broadly investigated the indirect impacts of the pandemic on non-COVID outcomes and reported ethnic differences. However, these two only included data until January 2021 at the latest. Other studies investigated just one disease area such as dementia or diabetes and frequently did not have the power to investigate specific ethnic groups.

*Added value of this study:* This is one of the largest studies to describe how the pandemic impacted ethnic differences in clinical monitoring at primary care and hospital admissions for non-COVID conditions (across four disease areas: cardiovascular disease, diabetes mellitus, respiratory disease and mental health) in England. A study population of nearly 15 million people, allowed the examination of five ethnic groups, and data until April 2022 allowed the evaluation of impacts for a longer period than previous studies.We showed that clinical monitoring had still not returned to pre-pandemic levels even by April 2022. Ethnic differences in clinical monitoring were seen pre-pandemic, though not in diabetes measures, these differences were either not impacted or reduced during the pandemic. We also showed that there were ethnic differences in hospital admissions, for many outcomes the pandemic did not impact these differences but there were some exceptions, in particular for diabetic ketoacidosis admissions in those of Black ethnicity and heart failure admissions for those of Black and Asian ethnicities.

*Implications of all the available evidence:* We found that the pandemic reduced ethnic inequalities for some outcomes (in hospitalisations for diabetic ketoacidosis and heart failure). However, these were driven by greater absolute increases in admissions for black and asian groups (diabetic ketoacidosis) and white groups (heart failure), which warrant further investigation to understand the underlying causes.

## Background

The COVID-19 pandemic directly impacted healthcare services across the world. This happened to different degrees in different countries, but had not recovered to pre-pandemic levels by the end of 2020.^1^ In the UK, primary care contacts and hospital admissions for a range of physical and mental health conditions decreased dramatically during 2020, most notably for anxiety, depression, Chronic Obstructive Pulmonary Disease (COPD) and cancer.^2,3^ COVID-19 disproportionately affects minority ethnic populations in the UK, with a higher risk of adverse COVID-19 outcomes, particularly in those of South Asian ethnicity.^4^ In addition to inequalities in the direct consequences of COVID-19, indirect healthcare consequences of the pandemic may also be unequal.^5^ In England, the pandemic impacted healthcare services differently across ethnic groups during 2020, with greater reductions in scheduled and unscheduled admissions, in those of non-White ethnicity compared with pre-pandemic.^3,6^ Differences have also been seen within specific disease areas.^7,8^ For example, there were increases in diabetic ketoacidosis (DKA) admissions for those of non-White ethnicity during the first wave of the pandemic.^8^ However, many studies have lacked sufficient power to compare specific ethnic groups, resulting in ‘White’ vs ‘non-White’ comparisons.^8^ Moreover, few studies have examined the impact of the pandemic on healthcare services beyond 2020.

Clinical monitoring refers to health measurements, such as blood pressure, that take place in primary care. These measures aim to prevent serious illness by identifying disease at an earlier stage and ensure known diseases are well managed.^9,10^ During the pandemic resources were diverted to COVID-19 related work, resulting in a reduction in monitoring.^11^ This reduction may be associated with poorer disease control and an increase in adverse outcomes and hospitalisation. Therefore, we chose to investigate clinical monitoring and hospitalisations within four disease areas (cardiovascular disease (CVD), diabetes mellitus (DM), respiratory disease and mental health), to align with previous work,^2^ and because these disease areas have evidence of ethnic differences in incidence and management.^12–16^ We aimed to determine the impact of the pandemic on ethnic differences in clinical monitoring and hospital admissions for non-COVID related conditions in England between 2020 and 2022.

## Methods

### Study design and data source

We conducted a population-based observational cohort study using OpenSAFELY-TPP, a data analytics platform created on behalf of NHS England to address urgent COVID-19 research questions (https://opensafely.org). Pseudonymised primary care electronic health records (EHR) from primary care software provider TPP, covering approximately 40%, and broadly representative of, the population of England,^17^ were linked to inpatient admissions data from the Hospital Episode Statistics for England Admitted Patient Care dataset (HES-APC) and mortality data from the Office for National Statistics (ONS). Data include pseudonymized data such as coded diagnoses, medications and physiological parameters. No free text data are included.

### Study population

The study included adults aged 18 years and over registered with a TPP practice between 1st March 2018 and 30th April 2022, with at least three months of registration prior to study entry (further detail: statistical analysis section). People were excluded if age, sex, geographic region, or Index of Multiple Deprivation (IMD) were missing, as missingness may indicate poor data quality. People were also excluded if their household size was greater than 15, to exclude people living in institutions, e.g. care home residents, who may have different clinical monitoring and hospital admissions patterns compared with the general population. People were followed from the start of the study period until the earliest of death, de-registration from the primary care practice, latest data availability, or the end of the study. Four disease-specific sub-populations were identified (described in the outcomes section).

### Study Measures

#### Exposures

The primary exposure was self-reported ethnicity defined using SNOMED CT morbidity codes in the primary care record. Where unavailable, information was supplemented with secondary care data.^18^ Ethnic groups were combined into the 2021 census categories, as follows: White (White British, White Irish, other White), Asian (Indian, Pakistani, Bangladeshi, other South Asian), Black (African, Caribbean, other Black), Mixed (White and Asian, White and African, White and Caribbean, other Mixed) and Other (Chinese, Arab, all others).

Ethnic differences in outcomes were compared: 1) before and after the introduction of lockdown in the UK on 23rd March 2020^19^ (defined as pre-pandemic and pandemic time); and 2) across six time periods during the pandemic (Figure 1).

**Figure 1:**
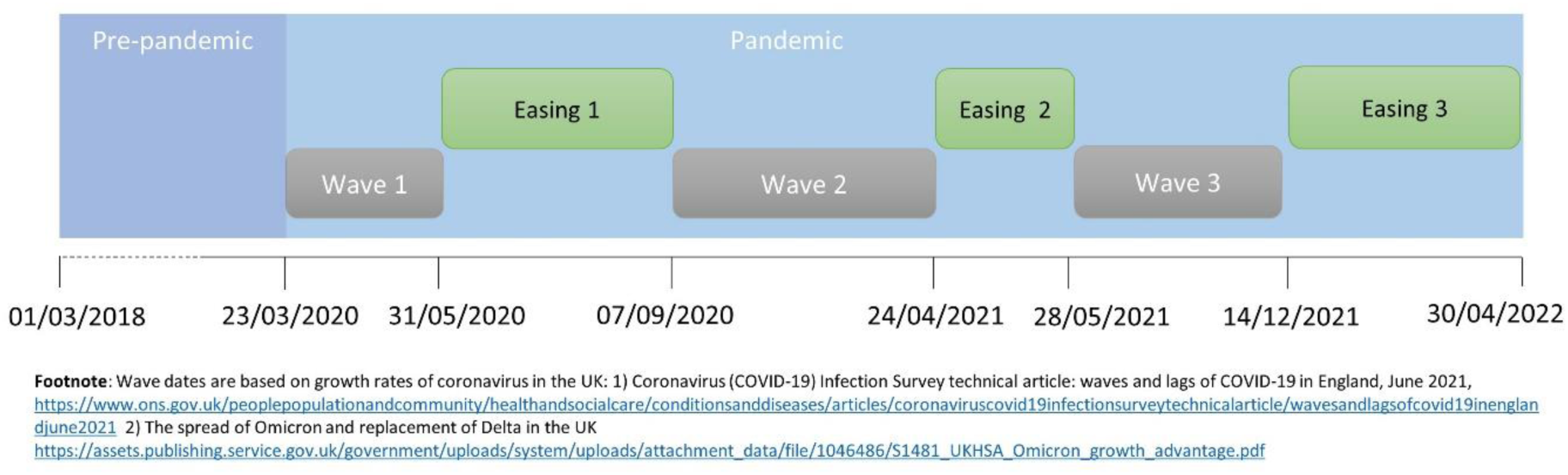
Study time-periods.

#### Outcomes

Study outcomes included clinical monitoring activities and hospital admissions related to four disease areas: DM, CVD, respiratory disease and mental health (Table 1).

**Table 1:** Outcome definitions and study populations

|  |  | Disease area |  |  |  |
| --- | --- | --- | --- | --- | --- |
|  |  | Diabetes | Cardiovascular disease | Respiratory disease | Mental health |
| **Clinical monitoring** | Outcomes measured | HbA1c, blood pressure | Blood pressure | COPD annual review<sup>1</sup>, asthma annual review<sup>2</sup> | Blood pressure |
|  | Study Population | People with Type 1 DM or Type 2 DM as coded on primary care record any time prior to study entry | People with coronary heart disease, history of stroke or TIA as coded in primary care record any time prior to study entry | <sup>1</sup> people aged >40 with a COPD code prior to study entry, <sup>2</sup> people with an asthma code in primary care record in the 3 years prior to study entry | People with schizophrenia, bipolar disorder and other psychoses as coded in primary care record any time prior to study entry |
| **Hospital admissions** | Outcomes measured | Admission with primary reason type 1 DM, type 2 DM or diabetic ketoacidosis | Admission with primary reason MI, stroke, heart failure or VTE | Admission with primary reason asthma exacerbation in those with asthma<sup>1</sup> or COPD exacerbation in those with COPD<sup>2</sup> | Admission with primary reason depression or anxiety |
|  | Study Population | People with type 1 DM or type 2 DM code in primary care record any time prior to study entry | General adult population | <sup>1</sup> age >40 with COPD code prior to study entry, <sup>2</sup> asthma code in primary care record in the 3 years prior to study entry | General adult population |

Footnote: Abbreviations: HbA1c: Haemoglobin A1c, COPD: chronic obstructive pulmonary disease, DM: diabetes mellitus, TIA: transient ischaemic attack, MI: myocardial infarction, VTE: venous thromboembolism.

#### Covariates

Demographic characteristics included age, sex, sustainability and transformation partnership (STP) region (NHS administrative geographical area), urban-rural classifier, deprivation, and shielding status. Deprivation was measured using quintiles of IMD based on a person’s postcode. People classed as extremely clinically vulnerable and therefore advised to shield were identified through SNOMED CT codes.^20^

### Statistical methods

The characteristics of the overall cohort on 1st January 2019, 2020 and 2021 were described by ethnic group. Two methods were used to estimate the impact of the pandemic on ethnic differences in outcomes: 1) interrupted time-series analysis; and 2) survival analysis.

#### Clinical monitoring

We calculated monthly crude rates of each clinical monitoring outcome, stratified by ethnicity. To measure monthly crude rates, study eligibility was assessed each month and individuals were included in the denominator for the whole month if they were eligible on the first of the month. Outcomes were counted once each month, but people could appear in multiple months if they had repeated records of the outcome in different months.

#### Pandemic impact on ethnic differences in clinical monitoring: interrupted time-series analysis

Monthly rates of each outcome were modelled in an ordinary least squares regression model with Newey-West heteroskedasticity-consistent standard errors and one lag to account for autocorrelation.^21–23^ The interruption was set at 23rd March 2020 (introduction of lockdown restrictions). To account for seasonal variation in outcome rates, season was included in the model as a four-level categorical variable: March-May, June-August, September-November, December-February. The rates were modelled with an interaction term for ethnicity to assess whether the ethnic patterning of outcomes changed from pre-pandemic to pandemic time.

Due to small numbers (n<10 per ethnic group in any single month) ethnic differences in hospital admissions could not be analysed using interrupted time-series analysis.

#### Hospital Admissions

We estimated hospital admission rates, by ethnic group, across eight pre-defined time periods (Figure 1). People who met the inclusion criteria at the start of each time-period were identified and followed from the start of the time-period until the earliest of death, de-registration from primary care practice, latest data availability, the end of the time-period, or until the first event for each hospital admission outcome.

#### Impact of pandemic on ethnic differences in hospital admissions

We used Cox proportional hazards regression to estimate ethnic differences in time to first hospital admission within each time period. We initially adjusted for age and sex, and then additionally for potential confounders: urban-rural classifier, deprivation, and shielding status. White ethnicity was the reference group and all models were clustered by STP region. To quantify the relative difference for each ethnic group, the ratio of HRs was calculated as the HR for each pandemic time period divided by the pre-pandemic HR. A ratio below one indicates a lower relative hazard of admission during the pandemic compared with pre-pandemic, a ratio above one indicates a higher relative hazard of admission during the pandemic time period.

#### Software and Reproducibility

We used Python 3.8 for data management, and Stata 17 and R for analyses. All code is shared for review and re-use under open licences at GitHub.com/OpenSAFELY. Code for data management and analysis, as well as codelists is archived online at https://github.com/opensafely/covid-collateral-research.

### Patient and Public Involvement

We have a publicly available website https://opensafely.org/ where we invite individuals to contact us regarding this study or the broader OpenSAFELY project.

## Results

As of 1st January 2020, 16,053,268 people met the inclusion criteria (Appendix). Ethnicity was missing for 1,122,912 (7%). Of those with known ethnicity, 12,926,485 (86.6%) were White, 1,096,398 (7.3%) were Asian, 381,441 (2.6%) were Black, 201,747 (1.4%) were of Mixed ethnicity, and 324,285 (2.2%) were of Other ethnicities. Compared with all other ethnic groups, those of White ethnicity were older, and lived in less deprived and more rural locations (Table 2). CVD was the most common comorbidity in those of White ethnicity (11.4%) while Type 2 DM was the most common comorbidity in those of Asian (12.6%) and Black (9.1%) ethnicities. Characteristics as of 1st January 2019 and 2021 were similar (Appendix).

**Table 2:** Baseline characteristics on 1st January 2020 by ethnic group

|  |  | All | White | Asian | Black | Mixed | Other | Missing |
| --- | --- | --- | --- | --- | --- | --- | --- | --- |
|  |  | N=16,053,268 | N=12,926,485 | N=1,096,398 | N=381,441 | N=201,747 | N=324,285 | N=1,122,912 |
| Age category | 18 - 40 years | 5,773,317 (36.0%) | 4,227,311 (32.7%) | 546,311 (49.8%) | 169,643 (44.5%) | 117,724 (58.4%) | 185,369 (57.2%) | 526,959 (46.9%) |
|  | 41 - 60 years | 5,284,691 (32.9%) | 4,207,590 (32.6%) | 382,006 (34.8%) | 158,168 (41.5%) | 63,377 (31.4%) | 99,890 (30.8%) | 373,660 (33.3%) |
|  | 61 - 80 years | 3,983,115 (24.8%) | 3,554,397 (27.5%) | 142,755 (13.0%) | 43,560 (11.4%) | 17,691 (8.8%) | 33,639 (10.4%) | 191,073 (17.0%) |
|  | >80 years | 1,012,145 (6.3%) | 937,187 (7.3%) | 25,326 (2.3%) | 10,070 (2.6%) | 2,955 (1.5%) | 5,387 (1.7%) | 31,220 (2.8%) |
| Sex | Male | 7,921,166 (49.3%) | 6,210,432 (48.0%) | 558,938 (51.0%) | 188,610 (49.4%) | 96,550 (47.9%) | 163,616 (50.5%) | 703,020 (62.6%) |
| IMD | 1 (most deprived) | 3,174,183 (19.8%) | 2,320,768 (18.0%) | 356,411 (32.5%) | 155,272 (40.7%) | 58,165 (28.8%) | 82,342 (25.4%) | 201,225 (17.9%) |
|  | 2 | 3,232,330<br>(20.1%) | 2,478,666<br>(19.2%) | 309,455<br>(28.2%) | 98,646<br>(25.9%) | 47,107<br>(23.3%) | 79,919<br>(24.6%) | 218,537<br>(19.5%) |
|  | 3 | 3,475,747<br>(21.7%) | 2,848,543<br>(22.0%) | 212,185<br>(19.4%) | 64,707<br>(17.0%) | 40,553<br>(20.1%) | 67,290<br>(20.8%) | 242,469<br>(21.6%) |
|  | 4 | 3,240,603<br>(20.2%) | 2,755,299<br>(21.3%) | 126,438<br>(11.5%) | 38,687<br>(10.1%) | 31,583<br>(15.7%) | 53,188<br>(16.4%) | 235,408<br>(21.0%) |
|  | 5 (least<br>deprived) | 2,930,405<br>(18.3%) | 2,523,209<br>(19.5%) | 91,909 (8.4%) | 24,129 (6.3%) | 24,339<br>(12.1%) | 41,546<br>(12.8%) | 225,273<br>(20.1%) |
| Type 1<br>diabetes | Yes | 113,640<br>(0.7%) | 99,350<br>(0.8%) | 7,163<br>(0.7%) | 2,950<br>(0.8%) | 1,160<br>(0.6%) | 1,225<br>(0.4%) | 1,792<br>(0.2%) |
| Type 2<br>diabetes | Yes | 1,158,128<br>(7.2%) | 928,618<br>(7.2%) | 138,022<br>(12.6%) | 34,800<br>(9.1%) | 10,831<br>(5.4%) | 15,239<br>(4.7%) | 30,618<br>(2.7%) |
| Asthma<sup>§</sup> | Yes | 1,399,693<br>(8.7%) | 1,215,301<br>(9.4%) | 83,180<br>(7.6%) | 25,166<br>(6.6%) | 16,664<br>(8.3%) | 13,551<br>(4.2%) | 45,831<br>(4.1%) |
| COPD<sup>%</sup> | Yes | 516,877<br>(3.2%) | 490,897<br>(3.8%) | 10,066<br>(0.9%) | 3,205<br>(0.8%) | 1,728<br>(0.9%) | 2,339<br>(0.7%) | 8,642<br>(0.8%) |
| CVD<sup>^</sup> | Yes | 1,835,952<br>(11.4%) | 1,686,486<br>(13.0%) | 69,528<br>(6.3%) | 21,660<br>(5.7%) | 9,170<br>(4.5%) | 12,608<br>(3.9%) | 36,500<br>(3.3%) |
| Serious<br>mental<br>illness<sup>*</sup> | Yes | 194,273<br>(1.2%) | 160,501<br>(1.2%) | 13,824<br>(1.3%) | 8,162<br>(2.1%) | 3,793<br>(1.9%) | 3,119<br>(1.0%) | 4,874<br>(0.4%) |

Footnote: <sup>§</sup> asthma code in primary care record in the 3 years prior to study entry, <sup>%</sup> age >40 with COPD code prior to study entry, <sup>^</sup> coronary heart disease, history of stroke or TIA as coded in primary care record any time prior to study entry, <sup>\*</sup>schizophrenia, bipolar disorder and other psychoses as coded in primary care record any time prior to study entry.

### Clinical monitoring

The monthly frequency of all clinical monitoring outcomes decreased after the start of the pandemic. The change between pre-pandemic and pandemic time was most pronounced for blood pressure monitoring across all disease areas and smallest for asthma annual reviews. Monitoring did not recover completely by April 2022 for most outcomes, with HbA1c monitoring recovering the most and asthma annual reviews remaining fairly constant at the lower rate (Figure 2).

**Figure 2:**
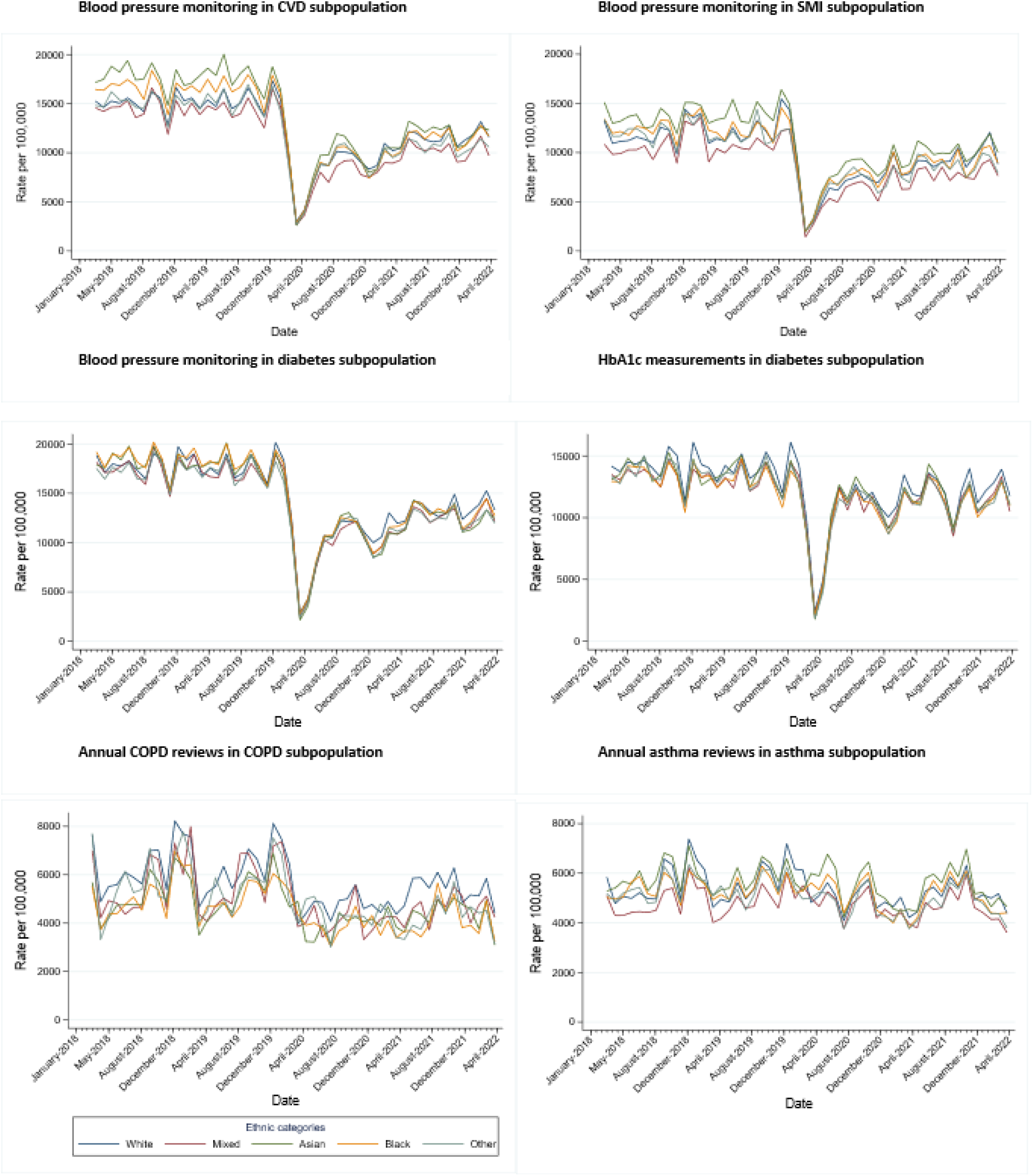
Monthly rates of clinical monitoring by ethnic group. Abbreviations: CVD: Cardiovascular disease, SMI: severe mental illness., COPD: chronic obstructive pulmonary disease.

#### Pandemic impact on ethnic differences in clinical monitoring

Interrupted time-series analysis indicated that, across the whole study period, ethnic differences in HbA1c monitoring were very small amongst people with diabetes. Amongst people with asthma and COPD, compared with people of White ethnicity, people of Mixed ethnicity received fewer asthma reviews and people of all minority ethnic groups received fewer COPD annual reviews. Blood pressure monitoring varied depending on the disease group. While blood pressure monitoring did not vary by ethnicity in those with diabetes, in those with CVD, blood pressure monitoring was lowest in those of Mixed ethnicity and highest in those of Asian and Black ethnicities. In those with serious mental illness, blood pressure monitoring was lowest in those of Mixed ethnicity and highest in those of Asian ethnicity (Figure 2).

Ethnic patterning of clinical monitoring remained unchanged between the pre-pandemic and pandemic periods for all outcomes, except for blood pressure monitoring in those with severe mental illness, where those of Asian ethnicity had fewer blood pressure measurements after the start of the pandemic (Figure 2).

### Rates of hospital admissions

Compared to pre-pandemic time, rates of hospital admissions increased for stroke and heart failure in the White ethnic group, with five additional admissions per month during the pandemic (stroke rate difference (RD) 5.2, heart failure RD: 5.4). For other ethnic groups and other CVD outcomes, differences in hospital admission rates were small (RD <=3).

Amongst people with diabetes, DKA admissions increased during the pandemic, most notably for people of Black ethnicity, with seven additional admissions per month (RD: 7.26). Hospital admissions for anxiety and depression were low for all ethnic groups (<1 per month) with less than one additional admission per month during the pandemic compared to pre-pandemic. Asthma and COPD hospital admissions decreased during the pandemic (Appendix).

### Pandemic impact on ethnic differences in hospital admissions

#### CVD

Prior to the pandemic, compared with those of White ethnicity, the age and sex adjusted hazard of stroke admission was higher in those of Black ethnicity. Hazards of VTE admission were lower in all minority ethnic groups compared with those of White ethnicity. Hazards of heart failure admission were higher in those of Black and Asian ethnicity, and hazards of MI related admissions were higher in those of Asian ethnicity compared to those of White ethnicity (Figure 3, Appendix).

**Figure 3:**
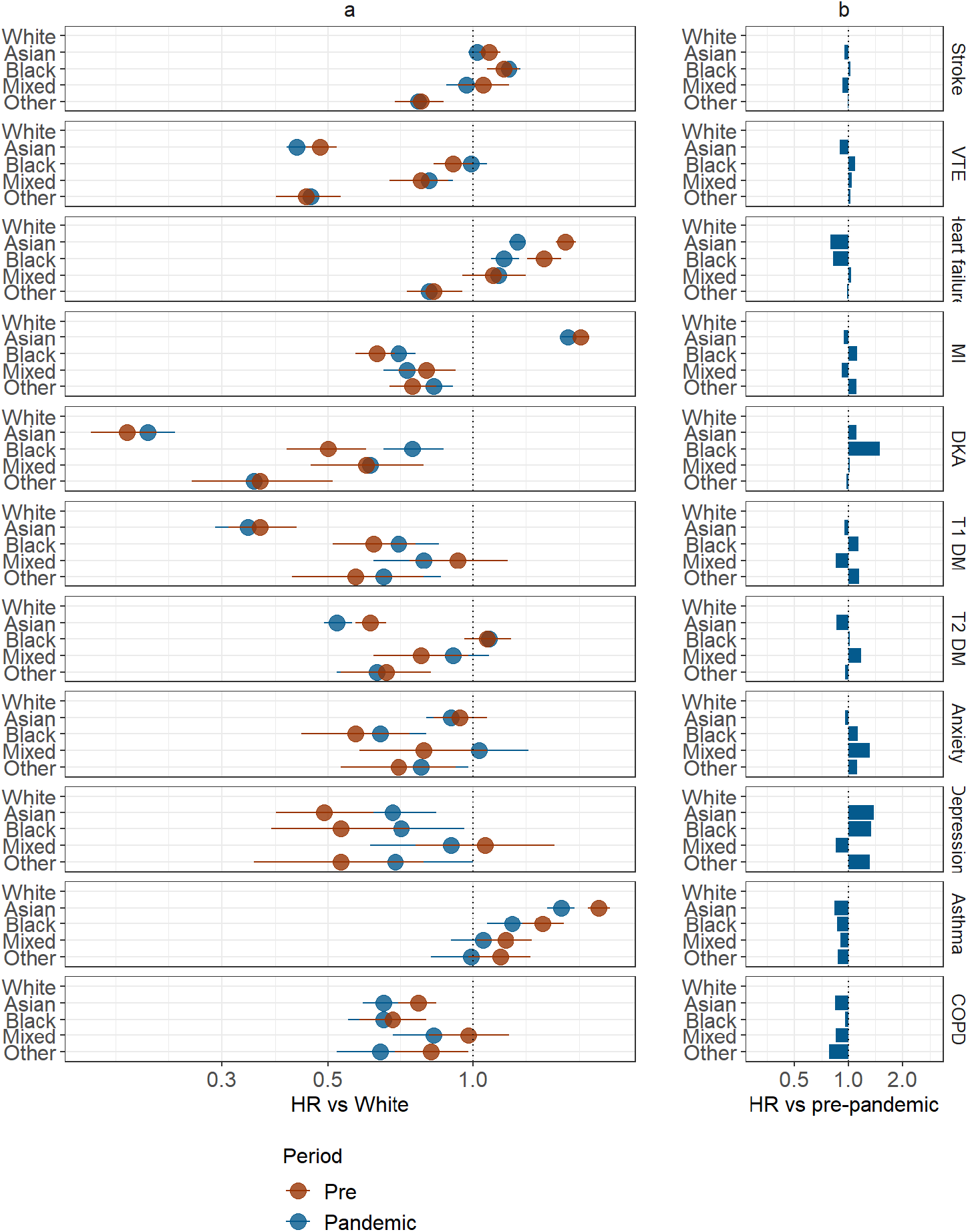
a) Age and sex adjusted hazard ratios pre-pandemic and pandemic for each non-White ethnic group versus White, b) Ratio of hazard ratios for each ethnic group.

While the ethnic patterning of hospital admissions for stroke, VTE, and MI remained unchanged between the pre-pandemic and pandemic periods, ethnic differences in heart failure admissions were attenuated during the pandemic in those of Asian (Pre-pandemic HR 1.56, 95% CI 1.49, 1.64, Pandemic HR 1.24, 95% CI 1.19, 1.29) and Black ethnicity (Pre-pandemic HR 1.41, 95% CI: 1.30, 1.53, Pandemic HR: 1.16, 95% CI 1.09, 1.25) (Figure 3, Appendix).

When comparing across wave and easing periods, ethnic differences for stroke and VTE remained small and consistent across the periods. The relative hazard of heart failure admission was lower for those of Black and Asian ethnicity in all pandemic waves compared with pre-pandemic. The hazard of MI admission in those of Black ethnicity was lower in Wave 1 compared with the pre-pandemic period and higher than pre-pandemic in all other waves (Appendix).

#### Diabetes

In age and sex adjusted analysis, all ethnic groups had a lower hazard of each outcome prior to and during the pandemic compared with White ethnicity. For DKA, the hazard of admission was higher during the pandemic for those of Black ethnicity (Pre-pandemic HR: 0.50, 95% CI 0.41, 0.60, Pandemic HR: 0.75, 95% CI: 0.65, 0.87), meaning differences between Black and White groups were attenuated during the pandemic. For Type 2 DM, the hazard of admission decreased for those of Asian ethnicity (Pre-pandemic HR 0.61, 95% CI 0.57, 0.66, Pandemic HR 0.52, 95% CI 0.49, 0.56), meaning differences between Asian and White groups widened.

When the pandemic period was split into wave and easing periods, the increase in DKA admissions in those of Black ethnicity was seen across all waves. There was a relative decline in admissions for Type 1 and Type 2 DM in those of Asian ethnicity across Waves 1 (23rd March 2020-30 May 2020), easing 1 (31 May 2020-6 September 2020), and Wave 2 (7 September 2020-23 April 2021) in particular.

#### Respiratory

For COPD, age and sex adjusted hazard of admission was lower in all ethnic groups compared with White before and during the pandemic. Ethnic differences widened during the pandemic, most notably for Asian groups relative to White (Pre-pandemic HR 0.77, 95% CI 0.70, 0.84, pandemic HR 0.65, 95% CI 0.59, 0.71).

For asthma, the Asian, Black and Mixed ethnic groups had a higher age and sex adjusted hazard of admission compared with those of White ethnicity prior to the pandemic. When comparing HRs between pre-pandemic and pandemic time periods, ethnic differences attenuated for all ethnic groups relative to those of White ethnicity, the biggest reduction was seen in those of Asian ethnicity (Pre-pandemic HR 1.83, 95% CI 1.74, 1.93, pandemic HR 1.53 (95% CI 1.43, 1.63)).

For both COPD and asthma admissions, the same patterns were seen across all pandemic time periods but differences reduced during later periods.

#### Mental health

All minority ethnic groups had lower or similar age and sex adjusted hazard of admission for both anxiety and depression, compared to those of White ethnicity, both before and during the pandemic. When comparing HRs between pre-pandemic and pandemic time periods for each ethnic group, differences between White and Mixed ethnicity were removed for anxiety related admissions during the pandemic (Pre-pandemic HR 0.79, 95% CI: 0.58, 1.07, Pandemic HR 1.03, 95% CI 0.81, 1.31). Ethnic differences in depression related admissions narrowed for those of Asian and Black relative to those of White ethnicity during the pandemic, (Asian ethnicity: Pre-pandemic HR 0.49, 95% CI 0.39, 0.62, Pandemic HR 0.68, 95% CI 0.55, 0.84, Black ethnicity: Pre-pandemic HR 0.53, 95% CI 0.38, 0.74, Pandemic HR 0.71, 95% CI 0.52, 0.96) (Figure 3, appendix).

For all hospital admission outcomes, additional adjustment for urban-rural classifier, deprivation, and shielding status made minimal difference to results (Appendix).

## Discussion

We found that, as of April 2022, primary care clinical monitoring across a range of conditions had still not returned to pre-pandemic levels. We saw ethnic differences in CVD, respiratory and mental health clinical monitoring, though there were some positive findings; ethnic differences in monitoring for people with diabetes were small, and although ethnic differences were apparent for other disease groups, these differences either remained the same, or were narrowed during the pandemic. In terms of hospital admissions, after accounting for age and sex, many ethnic differences remained unchanged during the pandemic, though there were some notable exceptions: ethnic differences attenuated during the pandemic for DKA admissions in the Black ethnic group and for heart failure admission for those of Asian and Black ethnicities relative to White. However, there were different mechanisms for these changes, for DKA admissions there was an absolute increase in rates in those of Black ethnicity that was not seen in other ethnic groups. For heart failure, there was an absolute increase in rates for all ethnic groups, but the biggest increase was seen in those of White ethnicity, narrowing relative differences.

Previous studies have shown that healthcare services in the UK, both broadly and within specific disease areas, were disrupted until the end of 2020.^2,3,24–28^ We show that disruption is still the case for clinical monitoring in 2022. We do not know if this is due to the health service being stretched ^29^ or people feeling reluctant to visit healthcare services, particularly if they are vulnerable. Although missed monitoring may represent appropriate reprioritisation of services,^30^ it may also represent missed opportunities for early diagnosis and prevention of serious outcomes. It is important to understand the characteristics of groups receiving less frequent care to determine whether these groups require targeted intervention. For example, in those with diabetes, reductions in routine diabetes monitoring have been associated with excess diabetes-related mortality.^31^

We showed that those of Black ethnicity had lower hazards of DKA admissions during both periods relative to those of White ethnicity, however the HR attenuated during the pandemic, indicating higher hazard of admission during the pandemic both in relative and absolute terms. Previous studies had shown ethnic differences in admissions for DKA in 2020, with increased admissions in non-White ethnic groups in the UK and non-Hispanic Black ethnicities in the US.^8,32^ We have not explored the reasons for the increase seen, although other studies have suggested it could be due to COVID-19 infection, which is known to disproportionately affect ethnic minorities, or worsening glycaemic control due to social restrictions.^33^

For those of Asian and Black ethnicities, we saw that the relative hazard of heart failure admission was attenuated during the pandemic, indicating a lower hazard of admission. Other studies in the general population have shown reductions in heart failure admissions during the pandemic (until mid-2020) compared to pre-pandemic.^34,35^ We saw similar heart failure admission rates during Wave 1 compared to pre-pandemic. Reductions in heart failure admissions could represent missed opportunities for preventive care, as studies have found increases in heart failure mortality alongside decreases in admissions for heart failure, particularly where there were large reductions in admissions.^34,36^ Similar reductions in admissions due to increased mortality are possible for all outcomes where admissions were reduced during the pandemic. Alternatively, reductions could indicate lower severity of these conditions, or these individuals may have been admitted with COVID as a primary diagnosis.

There was a higher hazard of admission for depression in those of Asian and Black ethnicities, compared with White ethnicity, although in absolute terms the number of additional admissions was small. Studies using data from a household survey in the UK in April 2020 found the highest levels of psychological distress in those of Asian ethnicity.^37,38^ It is possible that this translated to hospital admissions later in the pandemic. In addition, COVID-19 itself has been associated with mental health symptoms.^39^ As COVID-19 disproportionately affects ethnic minorities this could explain the increase in admissions relative to those of White ethnicity. Hospital admissions data only capture the most serious mental health cases, therefore exploration of other types of data (such as primary care records, patient-reported and mental health services data) may provide more insight.

A strength of this study is that we could investigate hospital admissions during periods of strict and relaxed restrictions into 2022. Broadly we saw that admissions were lower during Wave 1, which would be expected given the tight restrictions and uncertainty at that time. Many of the ethnic differences that were seen were consistent across all waves, though incidence rates were often highest during periods of easing restrictions, particularly the second period of easing (April 2021-May 2021). Further strengths of this study were the study population size, with 16 million people included. This allowed us to identify differences between individual ethnic groups rather than combining all ethnic minorities into one group. There were limitations: we were reliant on coding to identify exposures and outcomes, therefore misclassification is possible. If specific ethnic groups were less likely to present at healthcare services there could be differential misclassification for outcomes, although this may have less effect on hospitalisation outcomes due to their serious nature. Ethnicity information was missing for a small proportion of the population, we also did not have the power to investigate subcategories within the five ethnic groups. This study was primarily descriptive, therefore we did not explore potential explanatory factors for the differences seen or the impact of COVID-19. We also examined clinical monitoring and hospitalisations separately therefore could not examine the influence of clinical monitoring adherence on hospitalisations.

The causal mechanisms of ethnic differences seen are likely to be complex and disease specific, including genetic risk factors, differential exposure and vulnerability to COVID-19 and potential inequalities in health seeking behaviour and access to healthcare.^5^ Further research into the causal mechanisms, within disease areas where ethnic differences have been seen, is warranted.

In conclusion, these results demonstrate a consistent patterning of ethnic differences in relation to primary care monitoring of chronic conditions and hospital admissions in England, that has persisted over the period of the COVID-19 pandemic. It is critical to understand the causes of some of the differences identified and whether they represent inequities in access to or quality of care.

## Data Availability

All data were linked, stored and analysed securely within the OpenSAFELY platform https://opensafely.org/. All code is shared openly for review and re-use under MIT open license (https://github.com/opensafely/covid-collateral-research).

## Contributors

REC, JT, DP, EH, HC, PB, JKQ, LT, SML & RM were involved in the development of the study. REC, JT, DP, EH, BZ, EPKP, BM, RME and RM had access to the data. REC, JT, DP, EH and RM were responsible for data management and statistical analysis. REC and RM wrote the first draft of the manuscript. All authors contributed to and approved the final manuscript, and accept responsibility to submit for publication.

## Ethical approval

This study was approved by the Health Research Authority (REC reference 20/LO/0651) and by the LSHTM Ethics Board (reference 21863).

